# Childhood and adolescent immunisation intentions post-COVID-19 in Lusaka, Zambia

**DOI:** 10.64898/2026.09.21.26363608

**Authors:** Mwiza Nyasa, Anjali Sharma, Andrew D. Kerkhoff, Dennis Ngosa, Bertha Shamoya, Kombatende Sikombe, Sandra S. Simbeza, Nelly Zulu, Elvin H. Geng, Jake M. Pry, Noelle Le Tourneau

**Affiliations:** Centre for Infectious Disease Research in Zambia, Lusaka, Zambia; London School of Hygiene and Tropical Medicine, London, United Kingdom; Zambart, Lusaka, Zambia; Division of HIV, Infectious Diseases and Global Medicine Zuckerberg San Francisco General Hospital and Trauma Centre, University of California San Francisco, San Francisco, California, United States of America; Division of Infectious Diseases, School of Medicine, Washington University in St. Louis, Saint Louis, MO, United States of America; Division of Epidemiology, School of Medicine, University of California, Davis, California, United States of America

**Keywords:** vaccine confidence, vaccine hesitancy, childhood immunisation, herd immunity, vaccine-preventable diseases, public health outcomes

## Abstract

**Background:** Since the COVID-19 pandemic, parental delays and refusals of well-established, safe, and effective childhood vaccinations have contributed to the resurgence of infectious diseases. This raises concerns that COVID-19 vaccine hesitancy may also extend to novel vaccines for children when introduced in the future. This study examined caregivers’, who included healthcare workers (HCWs), perceptions of novel vaccinations for children ages 0-17 years old and intentions to vaccinate children and adolescents ages 12-17 years against COVID-19.

**Methods:** We conducted a secondary data analysis of a larger cross-sectional survey administered to parents/caregivers of children ages 0-17 years in Lusaka, Zambia, between 13 November and 15 December 2023. Vaccination intention for COVID-19 was specifically focused on caregivers of adolescents ages 12-17 years, following COVID-19 vaccination guidelines during the study. Descriptive statistics were used to explore demographic characteristics, vaccination history, and caregivers’ COVID-19 vaccination status, along with their children’s routine childhood vaccine histories and perceptions of COVID-19 and novel vaccinations.

**Results:** Among 277 caregivers (66.8% women, median age: 35 years), 93.1% reported that their child had received all recommended routine childhood vaccinations. COVID-19 vaccine uptake was higher among HCWs (68.2% with ≥2 doses) than among community members (40.8%) and having ≥2 doses was associated with higher COVID-19 vaccine uptake among their adolescents ages 12-17 years. 58.0% of caregivers indicated that their 12-17 years old adolescents had received at least one COVID-19 vaccine dose. Overall intention to further vaccinate children against COVID-19, especially among those already unvaccinated, remained low, with 24.7% of caregivers reporting no intent to vaccinate their children. Protection from hospitalization and death from malaria, TB, HIV, pneumonia, influenza, and diarrheal diseases were the most cited reason (79.1%) for intention to vaccinate children with novel vaccines for these diseases. The most prominent reasons for reluctance towards future vaccines were the fear of potential side effects (45.5%) and insufficient knowledge about vaccination benefits and risks (31.8%). Caregivers most trusted the Ministry of Health (86.6%) and healthcare professionals (66.8%) as sources of vaccine information.

**Conclusions:** While routine childhood vaccine uptake remained high, acceptance of COVID-19 and future novel vaccines for children was lower and was influenced by caregivers’ own vaccination status. Targeted and transparent communication through trusted sources such as the Ministry of Health and healthcare professionals may be critical for addressing safety concerns and strengthening confidence in novel childhood vaccines.

## Introduction

Childhood vaccinations are one of history’s greatest public health wins, saving millions of lives every year. However, these gains are fragile. When a lack of confidence in vaccines grows, as was observed during the COVID-19 pandemic, immunization coverage rates drop, allowing vaccine-preventable diseases like measles and polio to return (1). In addition to disruptions in regular health services and diverted resources during the pandemic (2), various reasons such as fear of COVID-19 infection at facilities where vaccines are administered contributed to parents’ lack of confidence in the vaccine toward routine childhood vaccines (3). This lack of confidence persisted post-COVID-19, leading to a drop in routine childhood vaccine uptake and a rise in numbers of unvaccinated children worldwide (2). In 2024, an estimated 18 million infants compared to 13 million in 2023, did not receive the first dose of diphtheria, tetanus and pertussis (DPT)-containing vaccine (zero-dose), an important indicator for a child’s access to health system (4), with a 7% increase in zero-dose children in Zambia from 2019 to 2024(5). Additionally, the number of infants not completing routine vaccinations, defined as not receiving the third DTP dose by their first birthday, rose from 6 million to 25 million (6) despite ready availability through the Expanded Programme on Immunization (EPI) (7, 8). Parental vaccine hesitancy persisted beyond infancy with a study across 12 sub-Saharan African countries revealing that 21.2% parents of children aged 19 months to 6 years were hesitant towards routine childhood immunization (9). In Zambia, disruptions during the COVID-19 pandemic led to an increase in measles cases and deaths among young children (median age of 3 years) and a 2% increase in monthly measles mortality in the post-pandemic period (3). Thus, lack of confidence in COVID-19 vaccines threaten progress in infectious disease control and undermines advancements in widespread access to vaccines (10).

While childhood vaccine coverage in Zambia is historically high compared to Western countries(11), there is concern that lack of confidence in the novel COVID-19 vaccine may extend to other new and routine childhood immunizations. While COVID-19 vaccine programs initially prioritised adults, many countries expanded access to include children aged 12 years and older. However, some countries reported vaccine hesitancy among adults for themselves and for their children including due to fear of side effects, misleading information, and uncertainty of the effectiveness of the vaccine for their children (12). Suboptimal uptake has also been observed for other vaccines targeting adolescents. For example, nearly half (46.1%) of the 4,830 caregivers of girls aged 9–17 years enrolled in a cross-sectional study in Nigeria, reported that the girls under their care had not received the human papillomavirus (HPV) vaccine (13). These findings suggest that challenges associated with acceptance of newly introduced vaccines may extend beyond COVID-19 and affect future vaccination programmes targeting older children and adolescents.

Emerging evidence also suggests considerable lack of confidence among adolescents themselves. A multi-country study among 2662 adolescents aged 10-19 years in rural and urban Ghana, Nigeria, Ethiopia and Tanzania reported high levels of vaccine hesitancy, with 39.4% reporting no intention and 9.6% uncertain about getting vaccinated due to concerns regarding necessity, effectiveness and safety. Among those willing to be vaccinated, motivations included protecting themselves or their families followed by parental vaccine (13). Similarly, a qualitative study in South Africa and Nigeria among 15-24 years old adolescents found that fear, distrust of government, conspiracy theories and unhappiness towards perceived coercion contributed to negative attitudes, while protecting family members, social responsibility for herd immunity, and resuming normal activities such as travel, socialising and being unmasked motivated vaccine acceptance (14).

On 5^th^ of October 2022 the Ministry of Health in Zambia announced the launch of a COVID-19 vaccination campaign targeting adolescents 12 to 17. However, evidence on determinants of vaccine hesitancy among caregivers of adolescents in Zambia remains scarce as apart from HPV vaccines there are no other vaccines targeted at this age group, and the influence of parental COVID-19 vaccination status on children’s vaccination is understudied.

To address these gaps, we examined caregivers’ perceptions of novel vaccinations for children aged 0 to 17 years and intentions to vaccinate their adolescents aged 12 to 17 years against COVID-19 following COVID-19 vaccination guidelines at the time of the study. This sub-study was nested in a larger study examining the association of COVID-19 vaccination uptake and vaccine intention for newly introduced adult vaccines among community members and HCWs in Lusaka, Zambia. We included community perspectives as public health efforts targeting childhood immunization must account for caregivers’ underlying perceptions and leverage trusted information sources to deliver effective vaccine messaging (15). We also included healthcare workers (HCWs) because they represent a critical interface between caregivers and vaccine introduction, and their attitudes can strongly influence parental decisions regarding childhood vaccinations (16, 17). Examining HCWs’ perspectives on childhood vaccination, including their willingness to vaccinate their own children, may provide important insight into both the drivers of vaccine acceptance and potential barriers to uptake.

## Methods

### Study Design

We conducted this secondary analysis as part of a larger cross-sectional survey in Lusaka, Zambia (18). For this analysis we sought to assess caregiver attitudes and preferences about childhood vaccination among community members and HCWs. Community members were recruited from randomly selected households in four high-density, low-income urban communities with relatively low COVID-19 vaccination coverage and HCWs were recruited from ten healthcare facilities chosen for diversity in size and geographic location. All participants were enrolled between 13 November and 15 December 2023, which was after COVID-19 waves and disruptions in Zambia. COVID-19 vaccinations were officially launched in April 2021for adults (19) and December 2021 for children 12 to 17 years, followed by two campaigns in May and October 2022 to encourage parents to have their children vaccinated (20) and reach the 70% vaccination goal for eligible populations (21), respectively.

### Study Population and Recruitment Strategy

Participants were eligible if they were aged 18 years or older, resided in Lusaka Province, and HCWs were eligible if they provided direct clinical or community-based healthcare services. Parents or primary caregivers, henceforth referred to as caregivers, were defined as those reporting being responsible for caregiving for any children under 18 years old at the time of screening.

HCWS were purposively selected from 10 healthcare facilities including one university tertiary hospital, five first-level hospitals, three Urban Health Centres, and one Rural Health Centre to represent a diversity of sizes and geographic locations; additional detail on the sampling approach has been previously described (18, 22). Recruitment occurred across five healthcare facility departments: Mother and Child Health (MCH), Outpatient (OPD), TB, Adolescent, and ART clinics. More details on the recruitment strategies have been previously reported (23).

Non-HCW community members participants were recruited from 10 administrative zones subdivided into 10 neighbourhoods of approximately 100 households. We randomly selected one zone per community and applied systematic household sampling. Within neighbourhoods, the initial household was selected using the “spin-the-bottle” technique, followed by every 5th household to the right for a total of 10 households per neighbourhood and 100 households per zone. The first eligible and consenting household member was recruited. To ensure the communities were well represented, extra data collections efforts were carried out on weekends specifically to invite and recruit people, mostly men, who tend to work regular hours during the week.

### Ethical Considerations

The study protocol was approved by the National Health Research Ethics Board (NHRA 000002/17/08/2023) and the Institutional Review Board at Washington University in St. Louis (IRB#: 202308190). All participants provided written informed consent in their preferred language.

### Data Collection and Management

Trained Research Assistants (RAs) collected data electronically on tablets in the participant’s preferred language (English, Nyanja, or Bemba) using Sawtooth Software Lighthouse Studio Offline Surveys. The survey included sociodemographic information, child immunization history, knowledge, attitudes, and concerns about both current and novel adult and childhood vaccines, sources of vaccine information and trust in information channels, and intentions to vaccinate children with currently available and future vaccines. Participants received 100

Zambian Kwacha (ZMW; ~USD $4 at the time of the study) as reimbursement for their time. We implemented daily data quality checks to ensure accuracy with the research team addressing any discrepancies in the collected data compared to the survey logs and completeness of survey responses as previously reported (22).

## Analysis

We summarized demographic characteristics, vaccination status, among caregivers using descriptive statistics. We examined factors associated with caregiver intention to vaccinate children for COVID-19 and potential new vaccines, stratified by participant type (community, HCW) and the number of COVID-19 vaccine doses the caregiver had received (0, 1, or ≥2 doses). We compared community and HCW attitudes to identify key differences between groups and if so, where interventions should focus (e.g., community education or HCW training and support). This comparison matters particularly for HCWs because they are a primary trusted source, and their own beliefs, including willingness to vaccinate their children, shape credibility and strength of health recommendations. Continuous variables were compared using independent sample t-tests, and categorical variables were compared using chi-square tests. Beyond attitudes, we also compared actual COVID-19 vaccine uptake to see whether parents’ own vaccination behaviour and attitudes predict whether their children get vaccinated. We also explored reasons reported by caregivers for and against vaccination for their children. All analyses were conducted in Stata (16.1, StataCorp LLC, College Station, TX).

## Results

A total of 277 participants with children aged <18 years (211 community members and 66 HCWs) were included in the analysis, with women comprising 66.8% of this caregiver population and a median age of 35 years (IQR 29-44 years) (Table 1). Differences between community members and HCWs were observed in education and employment with community members more likely to have no education (20.9%) or some secondary schooling (35.1%), while HCWs predominately had post-secondary education (75.8%). Community members were more often unemployed (25.6%) or self-employed (32.7%) compared to HCWs (0.0% and 0.0%, respectively), whereas HCWs were mainly formal employees (65.2%) or volunteers (34.8%). Most participants were married or cohabiting (67.5%). Approximately 12.0% reported living with HIV in both groups (community members and HCWs).

**Table 1.**
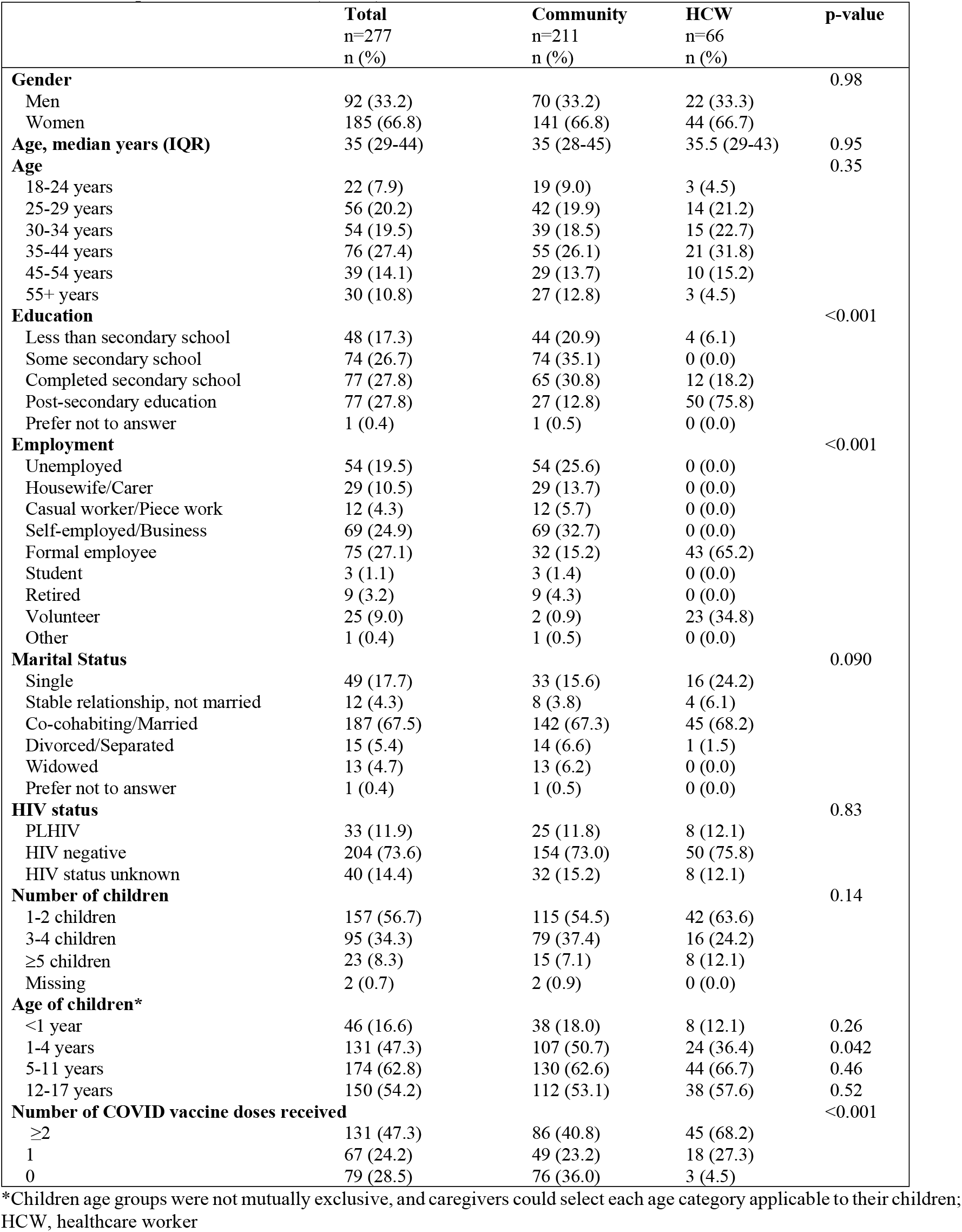
Participants characteristics (n=277)

|  | **Total**<br>n=277<br>n (%) | **Community**<br>n=211<br>n (%) | **HCW**<br>n=66<br>n (%) | **p-value** |
| --- | --- | --- | --- | --- |
| **Gender** |  |  |  | 0.98 |
| Men | 92 (33.2) | 70 (33.2) | 22 (33.3) |  |
| Women | 185 (66.8) | 141 (66.8) | 44 (66.7) |  |
| **Age, median years (IQR)** | 35 (29-44) | 35 (28-45) | 35.5 (29-43) | 0.95 |
| **Age** |  |  |  | 0.35 |
| 18-24 years | 22 (7.9) | 19 (9.0) | 3 (4.5) |  |
| 25-29 years | 56 (20.2) | 42 (19.9) | 14 (21.2) |  |
| 30-34 years | 54 (19.5) | 39 (18.5) | 15 (22.7) |  |
| 35-44 years | 76 (27.4) | 55 (26.1) | 21 (31.8) |  |
| 45-54 years | 39 (14.1) | 29 (13.7) | 10 (15.2) |  |
| 55+ years | 30 (10.8) | 27 (12.8) | 3 (4.5) |  |
| **Education** |  |  |  | <0.001 |
| Less than secondary school | 48 (17.3) | 44 (20.9) | 4 (6.1) |  |
| Some secondary school | 74 (26.7) | 74 (35.1) | 0 (0.0) |  |
| Completed secondary school | 77 (27.8) | 65 (30.8) | 12 (18.2) |  |
| Post-secondary education | 77 (27.8) | 27 (12.8) | 50 (75.8) |  |
| Prefer not to answer | 1 (0.4) | 1 (0.5) | 0 (0.0) |  |
| **Employment** |  |  |  | <0.001 |
| Unemployed | 54 (19.5) | 54 (25.6) | 0 (0.0) |  |
| Housewife/Carer | 29 (10.5) | 29 (13.7) | 0 (0.0) |  |
| Casual worker/Piece work | 12 (4.3) | 12 (5.7) | 0 (0.0) |  |
| Self-employed/Business | 69 (24.9) | 69 (32.7) | 0 (0.0) |  |
| Formal employee | 75 (27.1) | 32 (15.2) | 43 (65.2) |  |
| Student | 3 (1.1) | 3 (1.4) | 0 (0.0) |  |
| Retired | 9 (3.2) | 9 (4.3) | 0 (0.0) |  |
| Volunteer | 25 (9.0) | 2 (0.9) | 23 (34.8) |  |
| Other | 1 (0.4) | 1 (0.5) | 0 (0.0) |  |
| **Marital Status** |  |  |  | 0.090 |
| Single | 49 (17.7) | 33 (15.6) | 16 (24.2) |  |
| Stable relationship, not married | 12 (4.3) | 8 (3.8) | 4 (6.1) |  |
| Co-cohabiting/Married | 187 (67.5) | 142 (67.3) | 45 (68.2) |  |
| Divorced/Separated | 15 (5.4) | 14 (6.6) | 1 (1.5) |  |
| Widowed | 13 (4.7) | 13 (6.2) | 0 (0.0) |  |
| Prefer not to answer | 1 (0.4) | 1 (0.5) | 0 (0.0) |  |
| **HIV status** |  |  |  | 0.83 |
| PLHIV | 33 (11.9) | 25 (11.8) | 8 (12.1) |  |
| HIV negative | 204 (73.6) | 154 (73.0) | 50 (75.8) |  |
| HIV status unknown | 40 (14.4) | 32 (15.2) | 8 (12.1) |  |
| **Number of children** |  |  |  | 0.14 |
| 1-2 children | 157 (56.7) | 115 (54.5) | 42 (63.6) |  |
| 3-4 children | 95 (34.3) | 79 (37.4) | 16 (24.2) |  |
| ≥5 children | 23 (8.3) | 15 (7.1) | 8 (12.1) |  |
| Missing | 2 (0.7) | 2 (0.9) | 0 (0.0) |  |
| **Age of children\*** |  |  |  |  |
| <1 year | 46 (16.6) | 38 (18.0) | 8 (12.1) | 0.26 |
| 1-4 years | 131 (47.3) | 107 (50.7) | 24 (36.4) | 0.042 |
| 5-11 years | 174 (62.8) | 130 (62.6) | 44 (66.7) | 0.46 |
| 12-17 years | 150 (54.2) | 112 (53.1) | 38 (57.6) | 0.52 |
| **Number of COVID vaccine doses received** |  |  |  | <0.001 |
| ≥2 | 131 (47.3) | 86 (40.8) | 45 (68.2) |  |
| 1 | 67 (24.2) | 49 (23.2) | 18 (27.3) |  |
| 0 | 79 (28.5) | 76 (36.0) | 3 (4.5) |  |

\*Children age groups were not mutually exclusive, and caregivers could select each age category applicable to their children; HCW, healthcare worker

Most participants (56.7%) had one or two children, with 34.3% with three or four children, and 8.3% with five or more children. Among participants with children, 16.6% had at least one child aged <1 year, 47.3% had children aged 1–4 years, 62.8% had children aged 5–11 years, and 54.2% had children aged 12–17 years. Patterns were broadly similar across groups, except HCWs were less likely than community members to have young children aged 1–4 years (36.4% compared to 50.7%, respectively).

## General and EPI vaccination among children aged 0 to 17 years

Generally, caregivers indicated that vaccines were very important for their child’s health (93.1%), independent of their own vaccination status, with only 2.9% believing that vaccines were not important at all for their child’s health (Table S1). While there were differences in COVID-19 vaccination status for children by caregivers’ COVID-19 vaccination status, 93.1% of caregivers reported giving their children all recommended EPI vaccines, with no differences across caregivers’ COVID-19 vaccine history. Overall, 6.1% did not have their child receive a recommended EPI vaccine.

## COVID-19 vaccination among caregivers

Regarding COVID-19 vaccination, 68.2% of HCWs received two or more doses compared to 40.8% of community members, while 36.0% of community members remained unvaccinated for COVID-19 compared to only 4.5% of HCWs (Supplementary Table 1). Timing of caregivers’ first COVID-19 vaccine dose varied, with 37.9% vaccinated within 6 months of availability, 28.3% between 6–12 months, and 17.7% after 12 months. Caregivers with ≥2 doses (N=131; 47.3%) were more likely to vaccinate earlier than those with only 1 dose (42.7% vs. 28.4%, p=0.016). Vaccines were considered “very important” for children’s health by 93.1% overall, regardless of caregiver COVID-19 vaccination or healthcare worker status. Only 6.1% reported not vaccinating their child (zero-dose DPT) with a WHO recommended childhood vaccine (i.e., diphtheria, tetanus, pertussis, measles, polio, TB), with no differences across caregiver vaccination COVID-19 status or role.

## COVID-19 vaccination among adolescents aged 12 to 17

Among caregivers with adolescents aged 12 to 17 years (n=150), 58.0% indicated their child had received at least one COVID-19 vaccine dose and 38.0% reported their child had not received any COVID-19 vaccine doses. COVID-19 vaccine uptake was higher among children of caregivers with ≥2 COVID-19 doses (71.6%) compared with children of unvaccinated caregivers (36.6%, p=0.003), with no differences across HCW and community members (p=0.32) (Figure 1A, Table S1). Among all caregivers of adolescents, 24.7% did not intend to vaccinate their children for COVID-19, with 19.3% indicating they would vaccinate their children if required and 5.3% would never vaccinate their children. Among caregivers whose children were unvaccinated, only 24.6% intended to have their child vaccinated (15.8% in the next 6 months) (Figure 1B, Table S1).

**Figure 1.**
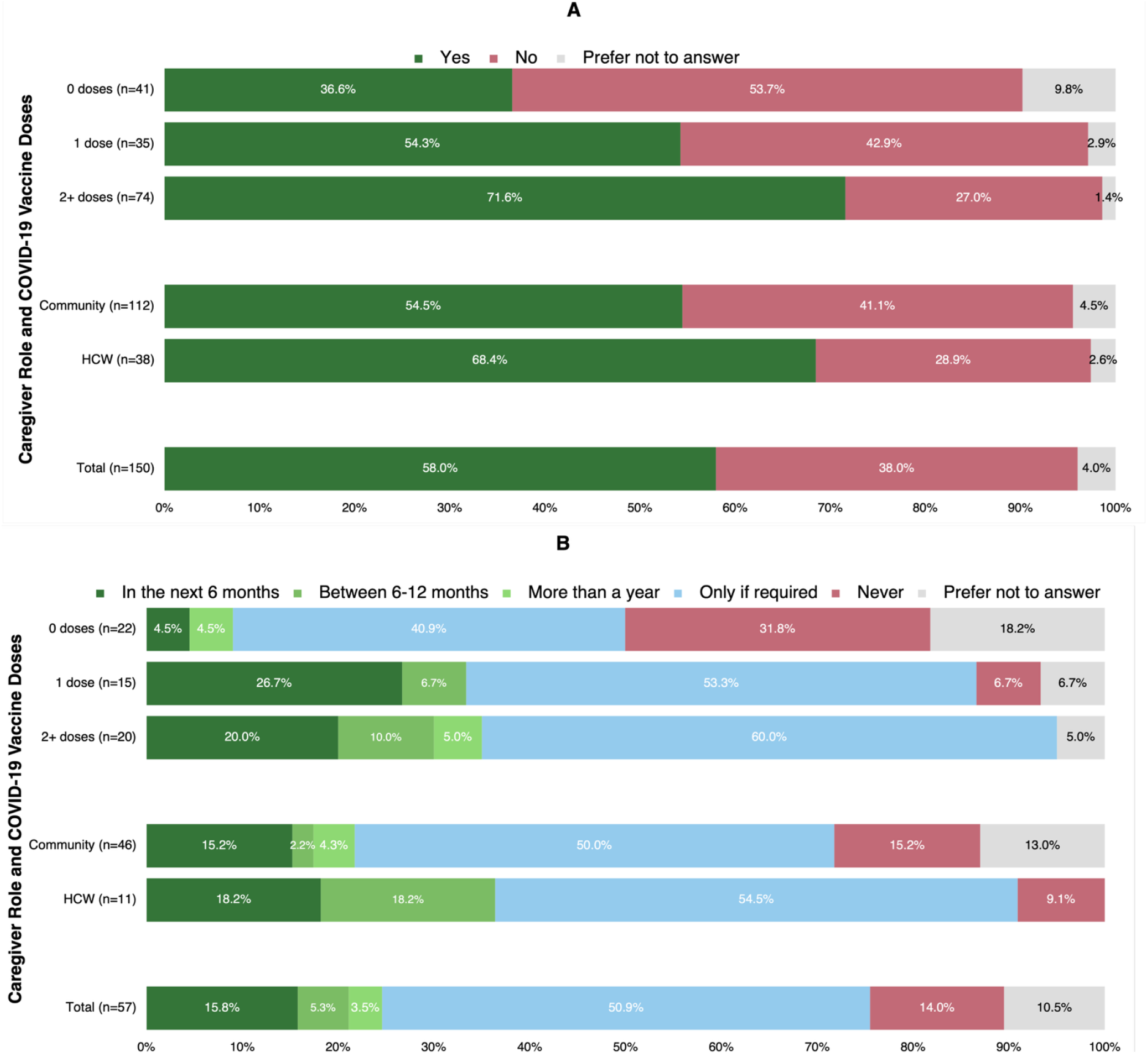
COVID-19 vaccination among adolescents aged 12-17 years. A) Any received COVID-19 doses among children 12-17 (n=150) and B) intention to vaccinate children 12-17 for COVID-19 among those not vaccinated (n=57), by caregiver COVID-19 vaccination and role

## Reasons for vaccinating among children aged 0 to 17 years

The most common reasons cited for vaccinating children against future diseases beyond COVID-19, including malaria, TB, HIV, pneumonia, influenza, and diarrheal disease, were protection from hospitalization or death (79.1%), protection against long-term effects of the disease (63.9%), and the general belief that vaccines are good for children (45.8%) (Figure 2A, Table S2A). Caregivers with ≥2 COVID-19 vaccine doses were more likely than unvaccinated caregivers to endorse vaccination for protection-related reasons, including hospitalization or death (82.4% vs. 68.4%, p=0.020), family protection (46.6% vs. 25.3%, p=0.006), potential economic/financial costs associated with the disease (38.9% vs 17.7%, p=0.005), and community protection (41.2% vs. 20.3%, p=0.002). HCWS were significantly more likely than community members to emphasize indirect benefits, including protecting family (59.1% vs. 30.3%, p<0.001), protecting the community (56.1% vs. 23.2%, p<0.001), and mitigating financial costs (51.5% vs. 23.7%, p<0.001). Few participants cited external pressures, such as government requirements (7.9%) or influence of family/friends (1.1%), and only 1.4% reported that vaccination was not relevant.

**Figure 2.**
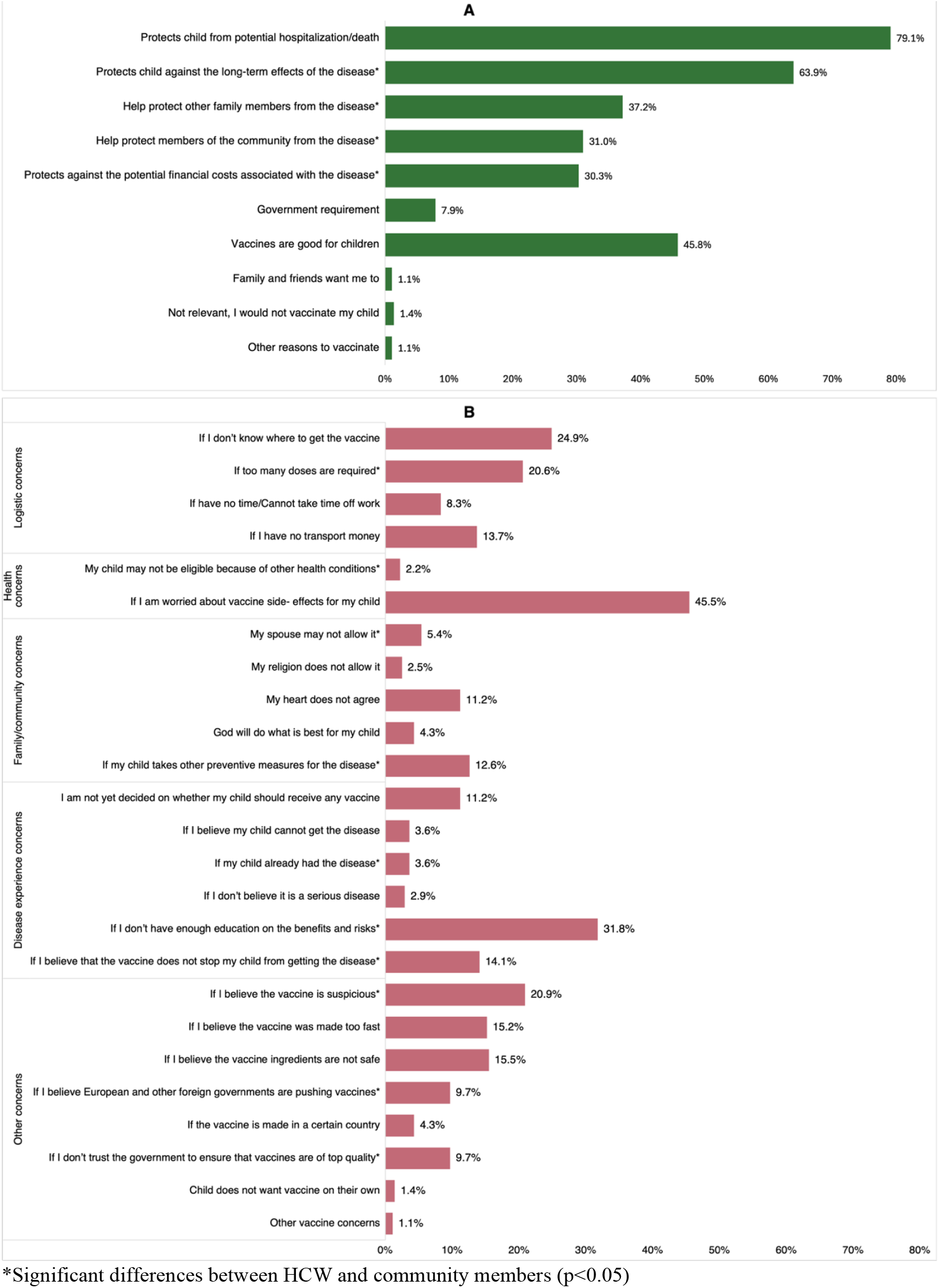
Hypothetical reasons for vaccinating or not vaccination children in the future against malaria, TB, HIV, pneumonia, influenza, updated/new COVID-19 booster, or diarrheal disease (n=277) A) Reasons for vaccinating children aged 0-17 years B) Reasons for not vaccinating children aged 0-17 years

## Reasons for not vaccinating among children aged 0 to 17 years

Caregivers reported a range of concerns about hypothetical vaccination for future or new vaccines (for their children, with variation by caregiver vaccine uptake and role (Figure 2B, Supplementary Table 2B). Health concerns were prominent, particularly fear of potential side effects (45.5%), reported more often by HCWs than community members (54.5% vs. 42.7%, p=0.090). Also, 31.8% reported insufficient knowledge about benefits and risks as a reason to not vaccinate, which was higher among HCWs than community members (42.4% vs. 28.4%, p=0.033). Logistic concerns included potential uncertainty about where to obtain the vaccine (24.9%), which was more common among caregivers with ≥2 doses compared to those unvaccinated (31.3% vs. 12.7%, p=0.009). Concerns about vaccine effectiveness and safety were also common: 14.1% believed vaccines would not prevent disease, more frequently among HCWs (25.8% vs. 10.4%, p=0.002), while 20.9% viewed vaccines as suspicious (31.8% vs. 17.5%, p=0.013). Distrust in government quality assurance was higher among HCWs (19.7% vs. 6.6%, p=0.002). Other concerns, such as beliefs about foreign influence (9.7%) or speed of development (15.2%), and safety of ingredients (15.5%) were also prevalent. While overall spousal disapproval of vaccination was an uncommon reason cited to not vaccinate (5.4%), HCWs were more likely to report spousal disapproval than community members (13.6% vs. 2.8%, p<0.001). Beliefs related to disease experience showed that caregivers whose child already had the disease were more likely to report this as a reason to not vaccinate (9.1% HCW vs. 1.9% community, p=0.006).

## Current and trusted sources of vaccination information

Caregivers reported multiple sources of vaccine information (median 4 sources, interquartile range [IQR]: 3-6 sources), with television (80.1%), health facilities (68.6%), radio (62.8%), and social media (57.0%) being the most cited overall (Supplementary Table 3). Use of information sources differed significantly by caregiver COVID-19 vaccine dose status and caregiver role (community versus HCW). Caregivers with no vaccine doses reported overall significantly (p<0.001) less vaccine information sources (median 3 sources, IQR: 2-5 sources) compared to those with 1 or ≥2 doses (median 5 sources, IQR 3-6 sources), including less use of health facilities, television, radio, print media, or workplaces as vaccine information sources. HCW caregivers were significantly (p<0.001) more likely than community caregivers to report more vaccine information sources (HCW: 6 sources, IQR: 5-8 sources) vs community: 4 sources, IQR: 2-5 sources) overall including social media, health facilities, workplaces, print media, schools and churches (Supplementary Table 3).

Trusted sources of vaccine information also varied by caregiver COVID-19 doses and caregiver role. Overall, the ministry of health (86.6%) and healthcare professionals (66.8%) were the most trusted vaccine information sources among caregivers (Table S3). Caregivers with any COVID-19 vaccine doses trusted more (p<0.001) sources (median: 2 sources, IQR: 1-4) than those with no doses (median: 1 source, IQR: 1-2). Caregivers with no COVID-19 vaccine doses were significantly (p=0.009) less likely to trust government sources (38.0%) compared to those with any doses (1 dose, 55.1%) and ≥2 doses (59.5%) but had no significant differences in trust in the ministry of health and healthcare professionals for vaccine information. HCWs trusted more sources than community caregivers (HCW: 3 (IQR 2-5) vs community: 2 (IQR 1-3) (p<0.001)), including government, healthcare professionals, scientists, international organizations and district health offices. Trust in friends, family, religious leaders, and community leaders was relatively low, but was more common among HCWS compared to community members. Trust in social media (10.1%), political leaders (2.2%) and traditional healers (1.1%) was low.

## Discussion

This secondary analysis among caregivers of children and adolescents in Lusaka, Zambia assessed attitudes and behaviours toward routine expanded programme on immunisation (EPI) and novel vaccines, including COVID-19 vaccination willingness for adolescents aged 12–17. Vaccine hesitancy, the reluctance or refusal to vaccinate despite availability of vaccines(24), is commonly understood through three interacting determinants: confidence, complacency, and convenience. Confidence, in particular, reflects trust in vaccine safety and effectiveness, in the health system and workers delivering them, and in the motives of policymakers who recommend them(25). While there were high uptake and perceived value of EPI vaccines, COVID-19 vaccination was low among adolescents 12 to 17, suggesting that caregiver hesitancy may be vaccine-specific rather than general, likely reflecting lingering uncertainty about newer vaccines. Notably, caregivers’ own COVID-19 vaccination status strongly predicted their children’s uptake, pointing to personal experience, rather than knowledge alone, as a key driver of novel vaccine decisions for children. Combined with caregivers’ trust in the Ministry of Health and healthcare professionals as vaccine information sources, these findings suggest that interventions to improve adolescent vaccine uptake should go beyond information to promote vaccine confidence among caregivers. Family-based strategies that engage already-vaccinated caregivers as peer champions, and that route messaging through the Ministry of Health and healthcare providers, may be more effective at building vaccine confidence and addressing hesitancy than generic education campaigns to influence uptake for future vaccination.

Caregivers considered EPI vaccines as widely important and recognized the importance of future vaccination of their children to protect them from hospitalization or death and the long-term effects of potential diseases. To our knowledge this is the first such data in our setting, and while comparability may be limited, similar trends have been noted across diverse settings. Similar reasons for vaccinating children were reported in a study in Myanmar, including wanting to keep children safe and less contagious (12). In contrast to our study where few participants (7.9%) cited external pressures (e.g., government requirements) as a reason to vaccinate, one study in the United States reported that top reasons for vaccination of children included being recommended by the child’s doctor and feeling pressure that the doctor would think less of the caregiver if their children were not vaccinated (18%) (26). Messaging around the importance of vaccination to prevent hospitalization, death, and long-term illness is essential for future childhood vaccine campaigns, where caregivers view this information as an important driver of vaccination for their children.

While EPI vaccination was reportedly high, consistent with another Zambian study in which 93% of parents reported willingness to vaccinate their children against cholera (27), there was hesitancy around COVID-19 vaccination for children, with caregiver COVID-19 vaccination status associated with COVID-19 vaccination uptake among adolescents aged 12 to 17. We found that COVID-19 vaccine uptake among 12- to 17-year-olds was relatively low, with only 56% reporting that their child had received a COVID-19 vaccine dose, and 25% of caregivers reporting that they did not intend to vaccinate their children against COVID-19. A study in five West African countries found similar results, where only 36% of caregivers said that they would vaccinate their children for COVID-19 if a vaccine were made available to them, while 25% said they would refuse to vaccinate their children (28). Similarly, trends have been noted in the US, where caregivers who had received the COVID-19 vaccine were more likely to have their children vaccinated than caregivers who had not received the vaccine themselves (29). Results from 8 Eastern Mediterranean countries also showed that children were 5 times more likely to be vaccinated when their caregivers were vaccinated than when they were not (30). During child health week events, campaigns have been used to target zero-dose children; one example is the Big Catch-up campaign in Zambia, which involved intensified outreach, community mobilization, and health service delivery (31). With nearly a quarter of caregivers hesitant to vaccinate children for COVID-19, we highlight the potential for targeted campaigns, such as the Big Catch-up, to inform caregivers about the importance of vaccinating children for future novel vaccines and to address primary concerns driving hesitancy.

As more vaccines are being produced, health communication strategies should be family-based and focus on providing caregivers with information on key issues of fear-based hesitancy identified, such as side effects, risks, and the benefits of vaccination with new vaccines, to both reduce parents’ vaccine-related fears and dispel misinformation. Vaccine hesitancy around fears of potential side effects was a highly reported concern by 55% of HCWs and 43% of community members. Similar findings in Myanmar showed that caregivers were hesitant to vaccinate their children as they felt that young children might not be fit to get vaccinated against COVID-19, as they were concerned about the perceived harmful effects of the vaccine (12). Concern about the speed at which the vaccine was developed was another reason given to not vaccinate (26). Clarifying the risks and benefits of side effects is crucial and requires sensitivity to parental concerns, as we know that parental vaccine status can influence their decision to vaccinate their children. Addressing parental concerns using events such as Child health week in Zambia can foster a positive perception of vaccines among caregivers, which, in turn, can influence their decision to vaccinate their children. Events such as these can provide caregiver sensitization that precedes vaccination by 3-6 months, allowing time to understand the risks and benefits and ensuring that trusted sources deliver vaccine messaging.

Leveraging trusted sources such as the Ministry of Health and health professionals to deliver these messages is crucial. Like our study, trust in HCWS to influence vaccine uptake was evident in a multi-country study done in Nepal, Senegal, and Zambia, where community health workers work with the community to share information, such as outreach and vaccine education (16). Doctors in the US were also seen as a trusted source of information, especially from family members who are doctors (32). Vaccinated HCWS, as trusted vaccine sources, can work with caregivers as community champions who can influence or encourage other caregivers to get their children vaccinated. Unvaccinated HCWS, on the other hand, can negatively influence the community by undermining vaccination campaigns through failing to recommend vaccines. That HCWs in our study reported concerns about novel vaccines more often than community members, despite their high endorsement of established EPI vaccines, complicates this champion role. It more likely reflects a reluctance to endorse a novel vaccine before the evidence is in than broad hesitancy, but whether that caution is reassuring or a scepticism that could reach the communities they serve, our data cannot resolve. Either way, HCW confidence in a new vaccine cannot be assumed and should be built ahead of rollout. Ministry of Health-branded messages and vaccinated HCWS as champions can be used to positively influence vaccine uptake in both unvaccinated HCWs and community members.

## Strengths and Limitations

Our study has several key strengths. The sample of both HCWs and community members provides insight into vaccination patterns and primary concerns for caregivers not only in the community but among HCWs who are trusted sources of vaccine information. The study has also provided valuable insights into the influence that caregivers can have on vaccine uptake among adolescents, offering a foundation for actionable health strategies to enhance immunizations through caregiver-targeted efforts.

Our study is not without limitations. While our sample population was recruited from ten health facilities and four diverse communities, all participants were from Lusaka, an urban city, with no rural representation, making our findings not generalizable to all parts of Zambia. The study relied on self-reporting of vaccination status among children and caregivers and may not reflect actual vaccine uptake. There could be potential social desirability bias to indicate more uptake of COVID-19 vaccine doses, however, the research assistants were experienced in collecting sensitive information and trained to build rapport with the participants in a way that encourages open and honest responses. Importantly, we recognize that adolescents themselves may have decision-making around their vaccination, and due to the nature of the study, we did not directly enrol or collect data from adolescents to explore their personal views on why they may or may not choose vaccination.

## Conclusion

Vaccine confidence among caregivers in this setting was specific rather than general, with established childhood vaccines remaining well accepted, and COVID-19 and future novel vaccines lower and shaped by caregivers’ COVID-19 vaccination behaviour. The task for future vaccine introduction is therefore to address the fear of side effects and the gaps in information on the risks and benefits highlighted by caregivers rather than confronting a broad hesitancy that our data do not show. Targeted campaigns aimed at caregivers in Zambia should leverage trusted sources, including the Ministry of Health and healthcare professionals, with clear information.

## Data Availability

A de-identified dataset supporting the conclusions of this article will be made publicly available.

## Acknowledgements

We sincerely thank all survey participants, the study teams who contributed to data collection, as well as the Johnson & Johnson Foundation, Scotland, whose support made this work possible.

## Supplementary Data

**Table S1.**
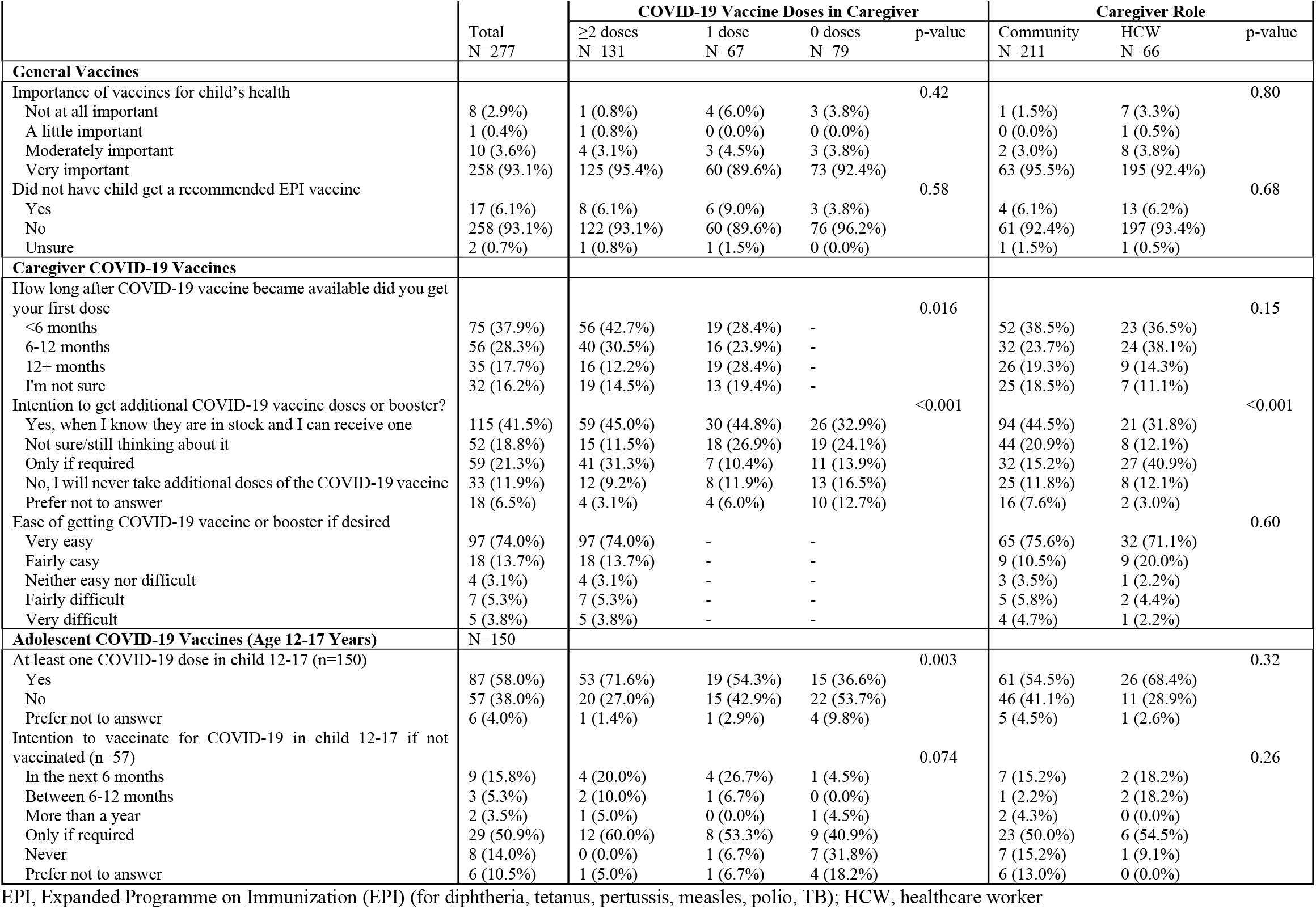
Vaccine history and intentions among caregivers and for children, by caregiver COVID-19 vaccination and role.

**Table S2A.** Hypothetical reasons for vaccinating children in the future against malaria, TB, HIV, pneumonia, influenza, updated/new COVID-19 booster, or diarrheal disease (n=277)

|  | Total<br>(n=277) | COVID-19 Vaccine Doses in Caregiver |  |  |  | Caregiver Role |  |  |
| --- | --- | --- | --- | --- | --- | --- | --- | --- |
|  |  | ≥2 doses<br>(n=131) | 1 dose<br>(n=67) | 0 doses<br>(n=79) | p-value | HCW<br>(n=66) | Community<br>(n=211) | p-value |
| Reasons for vaccinating child |  |  |  |  |  |  |  |  |
| Protects child from potential hospitalization/death | 219 (79.1%) | 108 (82.4%) | 57 (85.1%) | 54 (68.4%) | 0.020 | 56 (84.8%) | 163 (77.3%) | 0.19 |
| Protects child against the long-term effects of the disease | 177 (63.9%) | 94 (71.8%) | 34 (50.7%) | 49 (62.0%) | 0.013 | 51 (77.3%) | 126 (59.7%) | 0.010 |
| Help protect other family members from the disease | 103 (37.2%) | 61 (46.6%) | 22 (32.8%) | 20 (25.3%) | 0.006 | 39 (59.1%) | 64 (30.3%) | <0.001 |
| Help protect members of the community from the disease | 86 (31.0%) | 54 (41.2%) | 16 (23.9%) | 16 (20.3%) | 0.002 | 37 (56.1%) | 49 (23.2%) | <0.001 |
| Protects against potential economic/financial costs associated with the disease | 84 (30.3%) | 51 (38.9%) | 19 (28.4%) | 14 (17.7%) | 0.005 | 34 (51.5%) | 50 (23.7%) | <0.001 |
| Government requirement | 22 (7.9%) | 14 (10.7%) | 4 (6.0%) | 4 (5.1%) | 0.27 | 8 (12.1%) | 14 (6.6%) | 0.15 |
| Vaccines are good for children | 127 (45.8%) | 67 (51.1%) | 30 (44.8%) | 30 (38.0%) | 0.18 | 37 (56.1%) | 90 (42.7%) | 0.056 |
| Family and friends want me to | 3 (1.1%) | 3 (2.3%) | 0 (0.0%) | 0 (0.0%) | 0.18 | 1 (1.5%) | 2 (0.9%) | 0.70 |
| Other, specify: | 3 (1.1%) | 2 (1.5%) | 1 (1.5%) | 0 (0.0%) | 0.55 | 0 (0.0%) | 3 (1.4%) | 0.33 |
| I have vaccinated myself and want to protect my children the same way | 2 (0.7%) | 1 (0.4%) | 1 (0.4%) | 0 (0.0%) |  | 0 (0.0%) | 2 (0.7%) |  |
| Not relevant. I would not vaccinate my child | 4 (1.4%) | 0 (0.0%) | 1 (1.5%) | 3 (3.8%) | 0.082 | 1 (1.5%) | 3 (1.4%) | 0.96 |

**Table S2B.** Hypothetical reasons for not vaccinating children in the future against malaria, TB, HIV, pneumonia, influenza, updated/new COVID-19 booster, or diarrheal disease (n=277)

|  | Total (n=277) | COVID-19 Vaccine Doses in Caregiver |  |  |  | Caregiver Role |  |  |
| --- | --- | --- | --- | --- | --- | --- | --- | --- |
|  |  | ≥2 doses<br>(n=131) | 1 dose<br>(n=67) | 0 doses<br>(n=79) | p-value | HCW<br>(n=66) | Community<br>(n=211) | p-value |
| **Logistic concerns** |  |  |  |  |  |  |  |  |
| If I don't know where to get the vaccine | 69 (24.9%) | 41 (31.3%) | 18 (26.9%) | 10 (12.7%) | 0.009 | 16 (24.2%) | 53 (25.1%) | 0.89 |
| If too many doses are required | 57 (20.6%) | 28 (21.4%) | 11 (16.4%) | 18 (22.8%) | 0.61 | 25 (37.9%) | 32 (15.2%) | <0.001 |
| If have no time/Cannot take time off work | 23 (8.3%) | 15 (11.5%) | 5 (7.5%) | 3 (3.8%) | 0.14 | 5 (7.6%) | 18 (8.5%) | 0.81 |
| If I have no transport money | 38 (13.7%) | 22 (16.8%) | 8 (11.9%) | 8 (10.1%) | 0.35 | 11 (16.7%) | 27 (12.8%) | 0.43 |
| **Health concerns** |  |  |  |  |  |  |  |  |
| My child may not be eligible because of other health conditions (e.g., TB, sickle cell, heart condition, very sick, allergies) | 6 (2.2%) | 3 (2.3%) | 1 (1.5%) | 2 (2.5%) | 0.90 | 4 (6.1%) | 2 (0.9%) | 0.013 |
| If I am worried about vaccine side- effects for my child | 126 (45.5%) | 63 (48.1%) | 31 (46.3%) | 32 (40.5%) | 0.56 | 36 (54.5%) | 90 (42.7%) | 0.090 |
| **Family/community concerns** |  |  |  |  |  |  |  |  |
| My spouse may not allow it | 15 (5.4%) | 8 (6.1%) | 6 (9.0%) | 1 (1.3%) | 0.11 | 9 (13.6%) | 6 (2.8%) | <0.001 |
| My religion does not allow it | 7 (2.5%) | 5 (3.8%) | 1 (1.5%) | 1 (1.3%) | 0.43 | 3 (4.5%) | 4 (1.9%) | 0.23 |
| My heart does not agree | 31 (11.2%) | 12 (9.2%) | 7 (10.4%) | 12 (15.2%) | 0.40 | 9 (13.6%) | 22 (10.4%) | 0.47 |
| God will do what is best for my child | 12 (4.3%) | 5 (3.8%) | 4 (6.0%) | 3 (3.8%) | 0.75 | 4 (6.1%) | 8 (3.8%) | 0.43 |
| If my child takes other preventive measures for the disease | 35 (12.6%) | 21 (16.0%) | 5 (7.5%) | 9 (11.4%) | 0.21 | 13 (19.7%) | 22 (10.4%) | 0.048 |
| **Disease experience concerns** |  |  |  |  |  |  |  |  |
| I am not yet decided on whether my child should receive any vaccine | 31 (11.2%) | 14 (10.7%) | 8 (11.9%) | 9 (11.4%) | 0.96 | 9 (13.6%) | 22 (10.4%) | 0.47 |
| If I believe my child cannot get the disease | 10 (3.6%) | 4 (3.1%) | 3 (4.5%) | 3 (3.8%) | 0.87 | 3 (4.5%) | 7 (3.3%) | 0.64 |
| If my child already had the disease | 10 (3.6%) | 4 (3.1%) | 5 (7.5%) | 1 (1.3%) | 0.12 | 6 (9.1%) | 4 (1.9%) | 0.006 |
| If I don't believe it is a serious disease | 8 (2.9%) | 3 (2.3%) | 2 (3.0%) | 3 (3.8%) | 0.82 | 3 (4.5%) | 5 (2.4%) | 0.36 |
| If I don't have enough education on the benefits and risks | 88 (31.8%) | 49 (37.4%) | 18 (26.9%) | 21 (26.6%) | 0.16 | 28 (42.4%) | 60 (28.4%) | 0.033 |
| If I believe that the vaccine does not stop my child from getting the disease | 39 (14.1%) | 22 (16.8%) | 7 (10.4%) | 10 (12.7%) | 0.44 | 17 (25.8%) | 22 (10.4%) | 0.002 |
| **Other concerns** |  |  |  |  |  |  |  |  |
| If I believe the vaccine is suspicious | 58 (20.9%) | 27 (20.6%) | 11 (16.4%) | 20 (25.3%) | 0.42 | 21 (31.8%) | 37 (17.5%) | 0.013 |
| If I believe the vaccine was made too fast | 42 (15.2%) | 21 (16.0%) | 8 (11.9%) | 13 (16.5%) | 0.70 | 12 (18.2%) | 30 (14.2%) | 0.43 |
| If I believe the vaccine ingredients are not safe | 43 (15.5%) | 22 (16.8%) | 10 (14.9%) | 11 (13.9%) | 0.85 | 14 (21.2%) | 29 (13.7%) | 0.14 |
| If I believe European and other foreign governments are pushing vaccines | 27 (9.7%) | 12 (9.2%) | 7 (10.4%) | 8 (10.1%) | 0.95 | 13 (19.7%) | 14 (6.6%) | 0.002 |
| If the vaccine is made in a certain country | 12 (4.3%) | 8 (6.1%) | 2 (3.0%) | 2 (2.5%) | 0.39 | 4 (6.1%) | 8 (3.8%) | 0.43 |
| If I don't trust the government to ensure that vaccines are of top quality | 27 (9.7%) | 12 (9.2%) | 9 (13.4%) | 6 (7.6%) | 0.47 | 13 (19.7%) | 14 (6.6%) | 0.002 |
| Child does not want vaccine on their own | 4 (1.4%) | 0 (0.0%) | 1 (1.5%) | 3 (3.8%) | 0.082 | 0 (0.0%) | 4 (1.9%) | 0.26 |
| Other vaccine concerns | 3 (1.1%) | 1 (0.8%) | 1 (1.5%) | 1 (1.3%) | 0.88 | 0 (0.0%) | 3 (1.4%) | 0.33 |

**Table S3.**
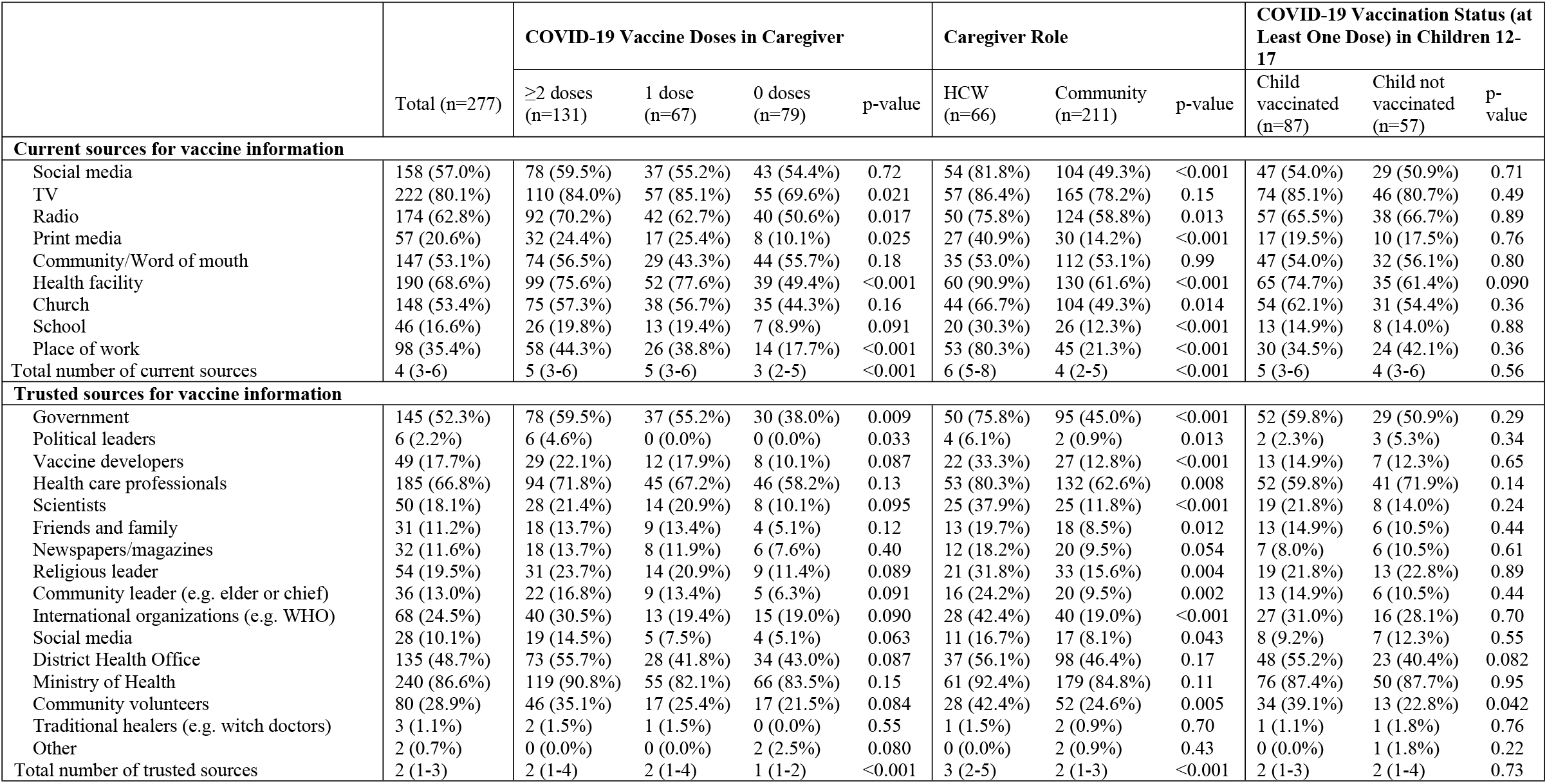
Current sources of vaccine-related information and trusted vaccine sources (n=277)

|  | Total (n=277) | COVID-19 Vaccine Doses in Caregiver |  |  |  | Caregiver Role |  |  | COVID-19 Vaccination Status (at Least One Dose) in Children 12-17 |  |  |
| --- | --- | --- | --- | --- | --- | --- | --- | --- | --- | --- | --- |
|  |  | ≥2 doses (n=131) | 1 dose (n=67) | 0 doses (n=79) | p-value | HCW (n=66) | Community (n=211) | p-value | Child vaccinated (n=87) | Child not vaccinated (n=57) | p-value |
| Current sources for vaccine information |  |  |  |  |  |  |  |  |  |  |  |
| Social media | 158 (57.0%) | 78 (59.5%) | 37 (55.2%) | 43 (54.4%) | 0.72 | 54 (81.8%) | 104 (49.3%) | <0.001 | 47 (54.0%) | 29 (50.9%) | 0.71 |
| TV | 222 (80.1%) | 110 (84.0%) | 57 (85.1%) | 55 (69.6%) | 0.021 | 57 (86.4%) | 165 (78.2%) | 0.15 | 74 (85.1%) | 46 (80.7%) | 0.49 |
| Radio | 174 (62.8%) | 92 (70.2%) | 42 (62.7%) | 40 (50.6%) | 0.017 | 50 (75.8%) | 124 (58.8%) | 0.013 | 57 (65.5%) | 38 (66.7%) | 0.89 |
| Print media | 57 (20.6%) | 32 (24.4%) | 17 (25.4%) | 8 (10.1%) | 0.025 | 27 (40.9%) | 30 (14.2%) | <0.001 | 17 (19.5%) | 10 (17.5%) | 0.76 |
| Community/Word of mouth | 147 (53.1%) | 74 (56.5%) | 29 (43.3%) | 44 (55.7%) | 0.18 | 35 (53.0%) | 112 (53.1%) | 0.99 | 47 (54.0%) | 32 (56.1%) | 0.80 |
| Health facility | 190 (68.6%) | 99 (75.6%) | 52 (77.6%) | 39 (49.4%) | <0.001 | 60 (90.9%) | 130 (61.6%) | <0.001 | 65 (74.7%) | 35 (61.4%) | 0.090 |
| Church | 148 (53.4%) | 75 (57.3%) | 38 (56.7%) | 35 (44.3%) | 0.16 | 44 (66.7%) | 104 (49.3%) | 0.014 | 54 (62.1%) | 31 (54.4%) | 0.36 |
| School | 46 (16.6%) | 26 (19.8%) | 13 (19.4%) | 7 (8.9%) | 0.091 | 20 (30.3%) | 26 (12.3%) | <0.001 | 13 (14.9%) | 8 (14.0%) | 0.88 |
| Place of work | 98 (35.4%) | 58 (44.3%) | 26 (38.8%) | 14 (17.7%) | <0.001 | 53 (80.3%) | 45 (21.3%) | <0.001 | 30 (34.5%) | 24 (42.1%) | 0.36 |
| Total number of current sources | 4 (3-6) | 5 (3-6) | 5 (3-6) | 3 (2-5) | <0.001 | 6 (5-8) | 4 (2-5) | <0.001 | 5 (3-6) | 4 (3-6) | 0.56 |
| Trusted sources for vaccine information |  |  |  |  |  |  |  |  |  |  |  |
| Government | 145 (52.3%) | 78 (59.5%) | 37 (55.2%) | 30 (38.0%) | 0.009 | 50 (75.8%) | 95 (45.0%) | <0.001 | 52 (59.8%) | 29 (50.9%) | 0.29 |
| Political leaders | 6 (2.2%) | 6 (4.6%) | 0 (0.0%) | 0 (0.0%) | 0.033 | 4 (6.1%) | 2 (0.9%) | 0.013 | 2 (2.3%) | 3 (5.3%) | 0.34 |
| Vaccine developers | 49 (17.7%) | 29 (22.1%) | 12 (17.9%) | 8 (10.1%) | 0.087 | 22 (33.3%) | 27 (12.8%) | <0.001 | 13 (14.9%) | 7 (12.3%) | 0.65 |
| Health care professionals | 185 (66.8%) | 94 (71.8%) | 45 (67.2%) | 46 (58.2%) | 0.13 | 53 (80.3%) | 132 (62.6%) | 0.008 | 52 (59.8%) | 41 (71.9%) | 0.14 |
| Scientists | 50 (18.1%) | 28 (21.4%) | 14 (20.9%) | 8 (10.1%) | 0.095 | 25 (37.9%) | 25 (11.8%) | <0.001 | 19 (21.8%) | 8 (14.0%) | 0.24 |
| Friends and family | 31 (11.2%) | 18 (13.7%) | 9 (13.4%) | 4 (5.1%) | 0.12 | 13 (19.7%) | 18 (8.5%) | 0.012 | 13 (14.9%) | 6 (10.5%) | 0.44 |
| Newspapers/magazines | 32 (11.6%) | 18 (13.7%) | 8 (11.9%) | 6 (7.6%) | 0.40 | 12 (18.2%) | 20 (9.5%) | 0.054 | 7 (8.0%) | 6 (10.5%) | 0.61 |
| Religious leader | 54 (19.5%) | 31 (23.7%) | 14 (20.9%) | 9 (11.4%) | 0.089 | 21 (31.8%) | 33 (15.6%) | 0.004 | 19 (21.8%) | 13 (22.8%) | 0.89 |
| Community leader (e.g. elder or chief) | 36 (13.0%) | 22 (16.8%) | 9 (13.4%) | 5 (6.3%) | 0.091 | 16 (24.2%) | 20 (9.5%) | 0.002 | 13 (14.9%) | 6 (10.5%) | 0.44 |
| International organizations (e.g. WHO) | 68 (24.5%) | 40 (30.5%) | 13 (19.4%) | 15 (19.0%) | 0.090 | 28 (42.4%) | 40 (19.0%) | <0.001 | 27 (31.0%) | 16 (28.1%) | 0.70 |
| Social media | 28 (10.1%) | 19 (14.5%) | 5 (7.5%) | 4 (5.1%) | 0.063 | 11 (16.7%) | 17 (8.1%) | 0.043 | 8 (9.2%) | 7 (12.3%) | 0.55 |
| District Health Office | 135 (48.7%) | 73 (55.7%) | 28 (41.8%) | 34 (43.0%) | 0.087 | 37 (56.1%) | 98 (46.4%) | 0.17 | 48 (55.2%) | 23 (40.4%) | 0.082 |
| Ministry of Health | 240 (86.6%) | 119 (90.8%) | 55 (82.1%) | 66 (83.5%) | 0.15 | 61 (92.4%) | 179 (84.8%) | 0.11 | 76 (87.4%) | 50 (87.7%) | 0.95 |
| Community volunteers | 80 (28.9%) | 46 (35.1%) | 17 (25.4%) | 17 (21.5%) | 0.084 | 28 (42.4%) | 52 (24.6%) | 0.005 | 34 (39.1%) | 13 (22.8%) | 0.042 |
| Traditional healers (e.g. witch doctors) | 3 (1.1%) | 2 (1.5%) | 1 (1.5%) | 0 (0.0%) | 0.55 | 1 (1.5%) | 2 (0.9%) | 0.70 | 1 (1.1%) | 1 (1.8%) | 0.76 |
| Other | 2 (0.7%) | 0 (0.0%) | 0 (0.0%) | 2 (2.5%) | 0.080 | 0 (0.0%) | 2 (0.9%) | 0.43 | 0 (0.0%) | 1 (1.8%) | 0.22 |
| Total number of trusted sources | 2 (1-3) | 2 (1-4) | 2 (1-4) | 1 (1-2) | <0.001 | 3 (2-5) | 2 (1-3) | <0.001 | 2 (1-3) | 2 (1-4) | 0.73 |

## References

1. UNICEF. Why vaccines matter for children. unicef for every child 2025.

2. UNICEF. New data indicates declining confidence in childhood vaccines of up to 44 percentage points in some countries during the COVID-19 pandemic. 2023.

3. Mwangilwa K, Sialubanje C, Chipoya M, Mulenga C, Mwale M, Chileshe C, et al. Attention to COVID 19 pandemic resulted in increased measles cases and deaths in Zambia. Tropical Medicine and Health. 2025;53(1).

4. Gavi. Gavi the vaccine alliance. 2025.

5. GAVI. Gavi Zambia:Country Information. 2025.

6. WHO. Global childhood vaccination coverage holds steady, 2025 [updated 2025–07–15. Available from: https://www.who.int/news/item/15-07-2025-global-childhood-vaccination-coverage-holds-steady-yet-over-14-million-infants-remain-unvaccinated-who-unicef.

7. Knisely JM, Erbelding E. Vaccines for Global Health: Progress and Challenges. The Journal of Infectious Diseases. 2025;232(1):25–7.

8. Shattock AJ, Johnson HC, Sim SY, Carter A, Lambach P, Hutubessy RCW, et al. Contribution of vaccination to improved survival and health: modelling 50 years of the Expanded Programme on Immunization. Lancet (London, England). 2024 May 25;403(10441).

9. Fadl N, Abdelmoneim SA, Gebreal A, Youssef N, Ghazy RM. Routine childhood immunization in Sub-Saharan Africa: addressing parental vaccine hesitancy. Public Health. 2024/01/01;226.

10. Cdc. Diseases that Vaccines Help Protect Against. 2025.

11. WHO U. Zambia: WHO and UNICEF estimates of immunization: 2025 revision. 2026.

12. Aung KT, Htun YM, Htet ZL, Soe YNM, Ko PK, Oo W, et al. Parental hesitancy on COVID-19 vaccination of children under the age of 16: A cross-sectional mixed-methods study among factory workers. PLOS ONE. 2025;20(6):e0327056–e.

13. Wang D, Chukwu A, Mwanyika-Sando M, Abubakari SW, Assefa N, Madzorera I, et al. COVID-19 vaccine hesitancy and its determinants among sub-Saharan African adolescents. PLOS Global Public Health. 2022;2(10):e0000611.

14. Casale M, Somefun O, Ronnie GH, Sumankuuro J, Akintola O, Sherr L, et al. Factors shaping Covid-19 vaccine acceptability among young people in South Africa and Nigeria: An exploratory qualitative study. PLOS Global Public Health. 2025;5(3):e0003795.

15. Oku A, Oyo-Ita A, Glenton C, Fretheim A, Ames H, Muloliwa A, et al. Perceptions and experiences of childhood vaccination communication strategies among caregivers and health workers in Nigeria: A qualitative study. PLOS ONE. 2017;12(11):e0186733.

16. Ogutu EA, Ellis AS, Hester KA, Rodriguez K, Sakas Z, Jaishwal C, et al. Success in vaccination programming through community health workers: a qualitative analysis of interviews and focus group discussions from Nepal, Senegal and Zambia. BMJ Open. 2024;14(4):e079358.

17. Biswas N, Mustapha T, Khubchandani J, Price JH. The Nature and Extent of COVID-19 Vaccination Hesitancy in Healthcare Workers. Journal of Community Health. 2021;46(6):1244–51.

18. Sharma A, Kerkhoff AD, Haambokoma M, Shamoya B, Sikombe K, Simbeza SS, et al. Intention to receive new vaccines post-COVID-19 pandemic among adults and health workers in Lusaka, Zambia. Vaccine. 2025;50:126846–.

19. WHO. Zambia launches the COVID-19 vaccination. WHO-ZAMBIA2021.

20. Cooperation MoFAaI. COVId 19 vaccination campaign.

21. UNICEF. The Government of the Republic of Zambia launches a new national COVID-19 vaccination campaign. UNICEF Zambi2022.

22. Tourneau NL, Sharma A, Pry JM, Haambokoma M, Shamoya B, Sikombe K, et al. Drivers of decision-making for future adult vaccines: a best–worst scaling among community members and health care workers in Zambia. Vaccine. 2026/01/05;70.

23. Sharma A, Kerkhoff AD, Haambokoma M, Shamoya B, Sikombe K, Simbeza SS, et al. Intention to receive new vaccines post-COVID-19 pandemic among adults and health workers in Lusaka, Zambia. Vaccine. 2025/03/19;50.

24. Sood Suruchi RF, Waaer Sarah, Block Suzanne. Guide for health workers on strengthening confidence in vaccines.; 2022.

25. Houle SKD, Andrew MK. RSV vaccination in older adults: Addressing vaccine hesitancy using the 3C model. Canadian Pharmacists Journal: CPJ. 2023 Nov 24;157(1).

26. Ruiz JB, Bell RA. Parental COVID-19 Vaccine Hesitancy in the United States. Public Health Reports. 2022;137(6):1162–9.

27. Pugliese-Garcia M, Heyerdahl LW, Mwamba C, Nkwemu S, Chilengi R, Demolis R, et al. Factors influencing vaccine acceptance and hesitancy in three informal settlements in Lusaka, Zambia. Vaccine. 2018/09/05;36(37).

28. Faye SLB, Krumkamp R, Doumbia S, Tounkara M, Strauss R, Ouedraogo HG, et al. Factors influencing hesitancy towards adult and child COVID-19 vaccines in rural and urban West Africa: a cross-sectional study. BMJ Open. 2022;12(4):e059138.

29. Byrne A, Thompson LA, Filipp SL, Ryan K. COVID-19 vaccine perceptions and hesitancy amongst parents of school-aged children during the pediatric vaccine rollout. Vaccine. 2022;40(46):6680–7.

30. Khatatbeh M, Albalas S, Khatatbeh H, Momani W, Melhem O, Al Omari O, et al. Children’s rates of COVID-19 vaccination as reported by parents, vaccine hesitancy, and determinants of COVID-19 vaccine uptake among children: a multi-country study from the Eastern Mediterranean Region. BMC Public Health. 2022;22(1).

31. Lungu V. Breaking Barriers:A Mother’s Journey to Embracing Child Health Services 2025 [Available from: https://www.unicef.org/zambia/stories/breaking-barriers.

32. Honcoop A, Roberts JR, Davis B, Pope C, Dawley E, Mcculloh RJ, et al. COVID-19 Vaccine Hesitancy Among Parents: A Qualitative Study. Pediatrics. 2023;152(5).

